# Rituximab counteracts loss of tolerance in membranous nephropathy patients through NK-mediated Treg induction

**DOI:** 10.1101/2025.10.10.25337710

**Authors:** Mounir El Maï, Maxime Teisseyre, Yousra Cheddadi, Vesna Brglez, Erendira Vazquez-Salazar, Sarah Nahon Carzo, Sami Addou, Kevin Zorzi, Marion Cremoni, Céline Fernandez, Barbara Seitz-Polski

**Affiliations:** Centre Hospitalier Universitaire de Nice, Centre de Référence Maladies Rares Syndrome Néphrotique Idiopathique et Glomérulonéphirte Extra-Membraneuse, Nice, France; Centre Hospitalier Universitaire de Nice, Laboratoire d’Immunologie et de Thérapie Cellulaire, Nice, France; Université Côte d’Azur, CNRS, INSERM, IRCAN, Nice, France

**Keywords:** Membranous nephropathy, Rituximab, Treg cells, NK cells, TGFβ

## Abstract

**Background:** Membranous nephropathy (MN) is a chronic, autoimmune kidney disease characterized by an autoimmune response against podocyte-specific antigens, compromising renal functions. Few studies attempted to mechanistically describe how rituximab influences Th cells and whether this impacts on clinical outcome.

**Methods:** Th cell profile of MN patients was characterized using peripheral blood mononuclear cell samples from a multi-centre, prospective clinical trial. Th cell subsets (Treg, Th17 and Th1) were measured by flow cytometry at baseline and after rituximab treatment during a two years follow-up. *In vitro* studies on fresh peripheral blood samples and isolated cell populations were performed to investigate the mechanisms through which rituximab induces Treg cells.

**Findings:** Before rituximab treatment, MN patients had a Th17/Treg imbalance when compared to age-matched healthy donors (p=0.004). Responders had significantly higher proportions of Treg cells post-rituximab than non-responders (p=0.037). *In vitro*, rituximab treatment induced Treg cells when using peripheral blood samples from MN patients, however the induction was lost when isolated T cells were used instead of whole blood (p=0.02). Rituximab did not directly induce Treg cells but required a specific cellular environment composed by at least B and Natural Killer (NK) cells as observed by treating pooled T, B and NK cells. Finally, using complementation assays, we demonstrated that rituximab-mediated Treg induction relied on cytokines, mainly Transforming growth factor β (TGFβ), secreted by activated NK cells.

**Interpretation:** Rituximab induces Treg through cytokines produced by NK cells probably during the mechanism of B cell depletion by antibody-dependent cell-mediated cytotoxicity. This mechanism could potentiate the likelihood of remission in MN patients by counteracting loss of immune tolerance.

## INTRODUCTION

Membranous nephropathy (MN) is a rare chronic autoimmune kidney disease and a leading cause of nephrotic syndrome in non-diabetic adults ^1–4^. It is characterized by an autoimmune response against podocyte-specific antigens, the phospholipase A2 receptor (PLA2R1) in the majority of cases (70-80%) and, less frequently, other antigens such as thrombospondin type-1 domain-containing 7A (THSD7A) observed in 3% of the cases^5–7^. This auto-immune reaction drives the deposits of auto-antibodies and complement factors leading to a thickening of the glomerular basement membrane^2,8^. The consecutive immune-mediated injury to glomerular podocytes results in proteinuria and most frequently to a nephrotic syndrome.

Rituximab, a monoclonal antibody targeting CD20 on B cells, has become a valuable treatment option for MN patients^9^. By depleting B cells and thereby reducing the production of pathogenic autoantibodies, rituximab leads to remission rates of 60–80% in MN patients^10–12^. However, the remaining 20-40% of patients do not respond to rituximab, even with significant B-cell depletion. This non-responsiveness has prompted further investigation into other immunological mechanisms in MN, particularly the involvement of T cells in sustaining disease activity and influencing therapeutic outcomes.

While B cells are a critical component in the autoimmune response, an imbalance in CD4 T helper (Th) cell differentiation also plays an essential role in the pathogenesis and progression of auto-immune diseases^13,14^. To coordinate various immune processes, CD4 Th cells differentiate into functional subtypes, including Th1, Th2, Th17, and regulatory T cells (Tregs), each defined by a distinct profile of cytokine produced. The Th1 pathway is associated with intracellular defense against infections while the pro-inflammatory Th17 pathway is involved in extracellular pathogen defense and involved in autoimmune diseases^15–17^. On the other hand, Tregs are essential for immune tolerance and to prevent excessive immune activation^13,18^. An imbalance of Th17/Treg ratio skewed toward Th17 response was reported in auto-immune diseases such as rheumatoid arthritis, systemic lupus erythematosus, multiple sclerosis, psoriasis, inflammatory bowel disease^13,14^. We previously described that MN patients had higher levels of pro-inflammatory cytokines of innate immunity (IL-6 and IL-1β), but also of Th17 (IL-17A), and Th2 (IL-4) pathways in peripheral blood immune cells following non-specific *in vitro* stimulation, compared to age- and sex-matched healthy controls. Notably, higher levels of IL-17A in MN patients were associated with increased risk of relapse and venous thromboembolic event. In contrast, Th1-(IFN-γ and IL-12p70) and Treg-(IL-10) type cytokines were lower in these patients^19^. Few studies attempted to describe an imbalance of Th cell differentiation in MN patients, especially of Th17/Treg imbalance^19–21^. Moreover, little is known about whether and how rituximab could reorient CD4 Th cell differentiation in MN patients^20^.

In this study, we performed a detailed characterization of Th cell subsets in MN patients. We further described long term effects of rituximab treatment on Th1, Th17 and Treg populations and investigated whether dynamic evolution of these lymphocyte subsets might predict the outcome of rituximab treatment in MN patients. Moreover, using *in vitro* experiments, we provided further mechanistic insights into how rituximab could favor a tolerogenic environment.

## METHODS

### Ethics and study participants

Patients samples used in this study were obtained from two distinct patient cohorts. First, frozen peripheral blood mononuclear cells (PBMCs) were obtained from a multi-centre, randomized, prospective clinical trial “Personalized Medicine for Membranous Nephropathy PMMN” registered under NCT03804359 (11-01-2019) and approved by the local ethics committee Sud-Ouest et Outre-Mer II^22^. From the 64 patients initially included in the study, 53 were chosen for this ancillary study on the basis of the treatment received during the entire duration of the study, i.e. 32 patients who received the standard GEMRITUX protocol (two 375 mg/m^2^ rituximab injections at one-week interval), and 21 patients who received two 1g rituximab injections at two weeks interval. The remaining 11 patients were excluded from this ancillary study since they either did not receive rituximab during the course of the study (N=8), due to loss to follow-up (N=2) or absence of medical insurance (N=1) (Supplemental figure 1). Patients were followed for two years after inclusion. The sample collected immediately before the first rituximab infusion was considered as baseline. Inclusion criteria for the PMMN cohort were: aged ≥18 years, anti-PLA2R1 activity detected by enzyme-linked immunosorbent assay (ELISA) or immunofluorescence assay, nephrotic syndrome defined by proteinuria > 3.5 g/24 h (or urine protein to creatinine ratio (UPCR) > 3.5 g/g) and serum albumin < 30 g/L, estimated Glomerular Filtration Rate (CKD-EPI formula) > 30 mL/min/1.73 m^2^, absence of immunosuppressive treatment in the last six months. The control group consisted of 15 age-matched healthy volunteers.

Second, fresh peripheral blood samples were obtained from a mono-center, prospective clinical trial BIOGEM registered under NCT05428605 (20-06-2022) and approved by the local ethics committee Sud-Est III. Inclusion criteria were: aged 18 years or more, anti-PLA2R1 or anti-THSD7A activity, relapsing MN patients with proteinuria > 3.5 g/24 h (or urine protein to creatinine ratio (UPCR) > 3.5 g/g), absence of immunosuppressive treatment in the last six months. All patients signed a written informed consent and the studies adhered to the declaration of Helsinki.

### Nuclear staining for Th subset characterization by flow cytometry

PBMCs isolated from PMMN cohort patients through Ficoll separation were first stained for surface markers for 30 minutes at 4°C in blocking solution (PBS, 5% FBS, 1% BSA, 2 mM EDTA) containing Brilliant stain buffer plus (BD Biosciences, Franklin Lakes, NJ) and fluorochrome-labeled monoclonal antibodies against CD45 (clone 2D1; RRID:AB_2814897; BD Biosciences); CD19 (clone SJ25C1; RRID:AB_1645728; BD Biosciences); CD3 (clone SK7; RRID:AB_2833003; BD Biosciences); CD4 (clone RPA-T4; RRID:AB_2744420; BD Biosciences); CD8 (clone SK1; RRID:AB_2868801; BD Biosciences); CD25 (clone 2A3; RRID:AB_2868687; BD Biosciences); and 7AAD (cat# 130-111-568; Miltenyi Biotech; Bergisch Gladbach, Germany). Then, after being washed, cells were fixed, permeabilized and stained using Foxp3/Transcription Factor Staining Buffer Kit (cat# TNB-0607-KIT; Tonbo biosciences, San Diego, CA, USA) according to manufacturer’s instructions. Nuclear staining was performed during 1h at room temperature using fluorochrome-labeled monoclonal antibody against FoxP3 (clone 259D/C7; RRID:AB_1645349; BD Biosciences), RORγt (clone 1181A; RRID:AB_3656845; Bio-Techne, Minnesota, USA) and T-bet (clone 04-46; RRID:AB_2738615; BD Biosciences). Samples were acquired and proportion of cell subsets were determined using BD FACSLyric flow cytometry system (BD Biosciences) as depicted in Supplemental figure 2.

### Intracellular staining for Th subset characterization by flow cytometry

One mL of peripheral blood from BIOGEM cohort patients was pretreated for 2h with rituximab (50 ng/mL) at room temperature prior to being transferred into QuantiFERON Monitor® tubes (Qiagen, Venlo, Netherlands) for 14h at 37°C. These tubes contained an anti-CD3 agonist antibody and Toll Like Receptor 7/8 agonist, enabling non-specific activation of T lymphocytes and innate immunity cells. GolgiStop solution (BD Biosciences) was then added and samples were left at 37°C for four additional hours. Red blood cells were depleted using PharM Lyse buffer (BD Biosciences). Samples were stained for surface markers for 30 minutes at 4°C in PBS containing Brilliant stain buffer plus and fluorochrome-labeled monoclonal antibodies against CD45 (clone HI30; RRID:AB_2716864; BD Biosciences); CD4 (clone RPA-T4; RRID:AB_2744420; BD Biosciences); CD8 (clone SK1; RRID:AB_1645736; BD Biosciences); CD127 (clone HIL-7R-M21; RRID:AB_1645548; BD Biosciences) and CD25 (clone 2A3; RRID:AB_2868687; BD Biosciences). Cells were fixed, permeabilized and stained using BD Cytofix/Cytoperm Plus kit (cat# 554715; BD Biosciences). Intracellular staining was performed using fluorochrome-labeled monoclonal antibodies against IL17A (clone N49-653; RRID:AB_2737902; BD Biosciences) and CD3 (clone OKT3; RRID:AB_2869862; BD Biosciences). Samples were acquired and proportion of cell subsets were determined using BD FACSLyric flow cytometry system (BD Biosciences) as depicted in Supplemental figure 3.

### ELISA

Plasma levels of IL-17A and IL-10 cytokines after non-specific stimulation in QuantiFERON Monitor tubes were measured for BIOGEM cohort using custom-designed microfluidic Simple Plex cartridge (Bio-Techne) and Ella plate reader (Bio-Techne), following the manufacturers’ instructions. All samples were measured diluted at 1:2.

### Treg and NK cells characterization by flow cytometry

One mL of peripheral blood samples from adult healthy donors were pretreated for 2h with rituximab (50 ng/mL) or obinutuzumab (50 ng/mL) at room temperature prior to non-specific stimulation of immune cells in QuantiFERON Monitor® tubes (Qiagen) for 18h at 37°C. Red blood cells were then depleted using Red blood lysis buffer (cat# 130-094-183; Miltenyi). Surface and nuclear staining were performed as described above after 40min incubation on ice in blocking solution and using fluorochrome-labeled monoclonal antibodies against CD45 (clone HI30; RRID:AB_2744405; BD Biosciences); CD19 (clone SJ25C1; RRID:AB_1645728; BD Biosciences); CD4 (clone RPA-T4; RRID:AB_2744420; BD Biosciences); CD25 (clone 2A3; RRID:AB_2868687; BD Biosciences); CD16a (clone 3G8; RRID:AB_2938676; BD Biosciences); CD56 (clone NCAM16.2; RRID:AB_2732054; BD Biosciences); CD3 (clone SK7; RRID:AB_2833003; BD Biosciences) and FoxP3 (clone 259D/C7; RRID:AB_1645349; BD Biosciences). Samples were acquired and proportion of cell subsets were determined using BD FACSLyric flow cytometry system (BD Biosciences) as depicted in Supplemental figure 4.

### Rituximab treatment on isolated cells

NK, T or B cells were isolated according to manufacturer’s instructions from five milliliters of peripheral blood samples from adult healthy donors using, respectively, MACSxpress Whole Blood NK Cell Isolation Kit (cat# 130-127-695), MACSprep™ HLA T (cat# 130-117-890) or B (cat# 130-110-130) Cell Isolation Kit (Miltenyi). One milliliter of each cell type eluate were used alone or combined (T; T+B or T+B+NK) and pretreated for 2h with rituximab (50 ng/mL) at room temperature prior to being transferred into QuantiFERON Monitor® tubes (Qiagen) for non-specific stimulation for 18h at 37°C. Surface and nuclear staining were performed as described above after 40min incubation on ice in blocking solution and using fluorochrome-labeled monoclonal antibody against CD45 (clone HI30; RRID:AB_2744405; BD Biosciences); CD19 (clone SJ25C1; RRID:AB_1645728; BD Biosciences); CD4 (clone RPA-T4; RRID:AB_2744420; BD Biosciences); CD25 (clone 2A3; RRID:AB_2868687; BD Biosciences); CD3 (clone SK7; RRID:AB_2833003; BD Biosciences) and FoxP3 (clone 259D/C7; RRID:AB_1645349; BD Biosciences).

### Complementation assay

NK, T and B cells were isolated from peripheral blood samples from adult healthy donors as described above. After 10 minutes centrifugation at 550g, B cells or B+NK cells were resuspended in one mL of RPMI containing 20% of respective donor plasma. Cells were pretreated for 2h with rituximab (50 ng/mL) at room temperature prior to being transferred into QuantiFERON Monitor® tubes (Qiagen) for non-specific stimulation for 18h at 37°C. Cells were centrifuged for 10min at 550g and 950µL of the complemented media were collected and treated or not for 2h at room temperature with galunisertib (10µM; Sigma; Massachusetts, USA) or emapalumab (10µg/mL; TargetMol, Boston, USA). Complemented media were then transferred onto donor respective T cells in a new QuantiFERON Monitor® tube (Qiagen). T cells were incubated for 18h at 37°C. Surface and nuclear staining were performed as described above using fluorochrome-labeled monoclonal antibody against CD45 (clone HI30; RRID:AB_2744405; BD Biosciences); CD19 (clone SJ25C1; RRID:AB_1645728; BD Biosciences); CD4 (clone RPA-T4; RRID:AB_2744420; BD Biosciences); CD25 (clone 2A3; RRID:AB_2868687; BD Biosciences); CD3 (clone SK7; RRID:AB_2833003; BD Biosciences) and FoxP3 (clone 259D/C7; RRID:AB_1645349; BD Biosciences).

### Statistics

Quantitative variables are reported as mean ± standard deviation or median [interquartile ranges] for, respectively, values that are normally distributed or not. For paired comparisons, a paired T-test was used for normally distributed data and Wilcoxon test for data that did not meet normality. Unpaired comparison of two groups was performed using a two-tailed Mann-Whitney tests. For multiple comparisons of paired data, an RM one-way analysis of variance (ANOVA) with Tukey’s post hoc correction was used for normally distributed data. A Chi-square test for trend was used to compare proportion distributions between groups. A critical value for significance of p < 0.05 was used throughout the study. All graphs and statistical analyses were performed with GraphPad Prism 8.4.3 (GraphPad Software Inc., San Diego, CA).

## RESULTS

### Patients and clinical data

The study enrolled 53 MN patients from the PMMN cohort^22^. Among MN patients, the majority were men (79%) and the average age was 59 years at inclusion. While 11 PMMN patients never entered clinical remission after rituximab treatment, 42 patients entered into clinical remission (which could be partial or complete with the criteria for both defined as UPCR < 3.5 g/g with a decrease greater than 50% from baseline, serum albumin >30 g/L and increase of serum creatinine lower than 20%). The majority (38) entered into clinical remission at last 12-month post-rituximab treatment while four reached clinical remission criterions at last at 18 months. We detected no difference between the two groups for all the demographic, except for age, and clinical parameters at diagnosis (Table 1).

**Table 1:** Baseline characteristics of the PMMN cohort.

|  | **All**<br>**(n=53)** | **REM+**<br>**(n=42)** | **REM-**<br>**(n=11)** | **p** |
| --- | --- | --- | --- | --- |
| Age (years) | 59.0 [55.0; 73.0] | 65.5 [56.0; 73.0] | 55.0 [48.0; 58.0] | **0.02** |
| Men (n, %) | 42 (79%) | 32 (76%) | 10 (91%) | 0.28 |
| Height (cm) | 173 ± 9 | 172 ± 9 | 176 ± 7 | 0.13 |
| Weight (kg) | 80.3 ± 14.8<sup>a</sup> | 78.8 ± 13.9<sup>a</sup> | 85.9 ± 17.5 | 0.23 |
| BMI (kg/m<sup>2</sup>) | 26.7 ± 4.2<sup>a</sup> | 26.5 ± 4.0<sup>a</sup> | 27.6 ± 4.8 | 0.52 |
| Systolic BP (mmHg) | 139 ± 18 | 139 ± 19 | 138 ± 14 | 0.82 |
| Diastolic BP (mmHg) | 86 [76; 90] | 82 [74; 90] | 89 [79; 95] | 0.18 |
| Creatinine (μmol/L) | 99.0 [80.5; 129.5] | 101.9 [80.8; 132.5] | 99.0 [79.0; 113.0] | 0.38 |
| Albumin (g/L) | 25.0 ± 4.0 | 25.2 ± 3.9 | 24.0 ± 4.8 | 0.43 |
| eGFR (mL/min/1.73 m<sup>2</sup>) | 67.3 ± 24.7 | 62.3 ± 24.8 | 74.9 ± 23.7 | 0.25 |
| UPCR (g/g) | 7.1 ± 2.6<sup>a</sup> | 7.1 ± 2.6 | 7.1 ± 2.6 | 0.98 |
| Edema | 27 (51%) | 19 (45%) | 8 (73%) | 0.10 |
| Anti-PLA2R1 (RU/mL) | 75 [31; 169]<sup>b</sup> | 78 [27; 161] | 91 [56; 419]<sup>a</sup> | 0.27 |
| Epitope spreading (n, %) | 27 (52%)<sup>a</sup> | 21 (50%) | 6 (60%)<sup>a</sup> | 0.57 |
| Time since the onset of symptoms (months) | 4.0 [2.4; 7.5] | 4.0 [2.4; 7.8] | 4.9 [1.9; 7.5] | 0.84 |
| History of MN (n, %) | First course: 34 (64%)<br>Relapse: 19 (36%) | First course: 26 (62%)<br>Relapse: 16 (38%) | First course: 8 (73%)<br>Relapse: 3 (27%) | 0.51 |
| Anti-hypertensive treatment: diuretics, beta blockers, calcium inhibitors (n, %) | None: 8 (15%)<br>1 drug: 27 (51%)<br>2 drugs: 12 (23%)<br>3 drugs: 6 (11%) | None: 6 (14%)<br>1 drug: 22 (52%)<br>2 drugs: 8 (19%)<br>3 drugs: 6 (14%) | None: 2 (18%)<br>1 drug: 5 (45%)<br>2 drugs: 4 (36%)<br>3 drugs: 0 (0%) | 0.41 |
| RAAS blockers (n, %) | None: 1 (2%)<br>1 drug: 49 (92%)<br>2 drugs: 3 (6%) | None: 1 (2%)<br>1 drug: 39 (93%)<br>2 drugs: 2 (5%) | None: 0 (0%)<br>1 drug: 10 (91%)<br>2 drugs: 1 (9%) | 0.76 |
| Other symptomatic treatment (n, %) | None: 49 (92%)<br>1 drug: 4 (8%) | None: 38 (90%)<br>1 drug: 4 (10%) | None: 11 (100%)<br>1 drug: 0 (0%) | 0.29 |

BP: blood pressure; eGFR: estimated glomerular filtration rate; BMI: body mass index; PLA2R1: phospholipase A2 receptor; MN: membranous nephropathy; UPCR: urine protein to creatinine ratio; RAAS: renin-angiotensin-aldosterone system
<sup>a</sup> Missing value for one patient
<sup>b</sup> Missing values for two patients
<sup>c</sup> Missing value for one patient without an immunological workup at month-0 was replaced with a previous immunological workup at screening one month before enrolment in the study. Of note, the epitope spreading result (i.e., no spreading) was identical pre-enrolment and at month-3 follow-up visit.

### T helper subsets dysregulation in MN at baseline

An imbalance in CD4 T helper (Th) cell differentiation has been highlighted in the development and outcome of auto-immune diseases^13,14^. In order to characterize the dysregulation in Th cell differentiation in MN, we first measured on the PMMN cohort the frequency of Treg, Th17 and Th1 cells at baseline by flow cytometry analyses using well described specific nuclear markers for each cell type. Interestingly, we observed lower levels of Treg (Foxp3+, CD25^high^ cells) among CD4+ cells in MN patients compared to HD (median 2.95% [2.04; 3.92] vs 4.89% [3.52; 5.92]; p=0.008; Figure 1A). In contrast, MN patients were enriched in Th17 (RORγt+ cells) among CD4+ cells compared to HD (median 1.55% [0.77; 2.80] vs 0.71% [0.51; 1.07]; p=0.014; Figure 1B). Accordingly, Th17/Treg balance ratio increased toward a pro-inflammatory profile in MN patients (median 0.46 [0.21; 0.97] vs 0.16 [0.13; 0.26]; p=0.004; Figure 1C). However, no difference was detected in Th1 levels (CD4+/T-bet+ cells) between MN patients and HD (median 1.06 [0.45; 2.81] vs 0.45 [0.26; 1.03]; p=0.12) but we observed higher Th1/Treg ratio in MN patients (median 0.42 [0.09; 0.85] vs 0.08 [0.07; 0.29]; p=0.04; Supplemental figure 5).

**Figure 1:**
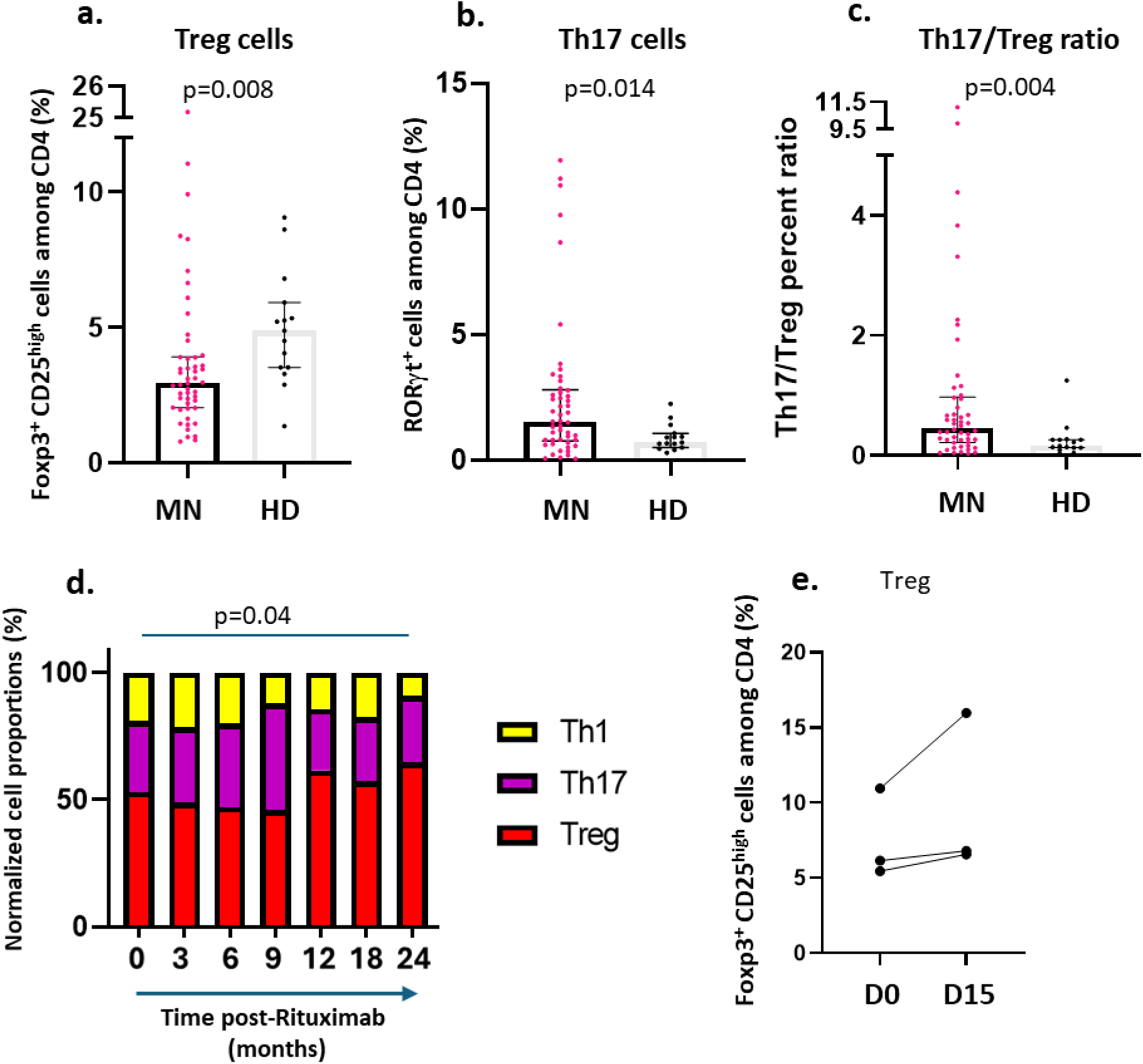
Dysregulation of Th17/Treg balance in MN patients. MN patients are characterized by reduced levels of Treg cells and higher levels of Th17 cells and Th17/Treg ratio. **a-c)** Flow cytometry quantification of Treg cell (Foxp3+, CD25high) **(a**), Th17 cells (RORgt+) **(b)** among peripheral CD4+ cells and Th17/Treg ratio **(c)** in MN patients from the PMMN cohort (MN; n=50) compared to healthy donors (HD; n=15). **d)** Kinetics of the relative proportion of Treg, Th17 and Th1 cells in MN patients (n=50) up to 24 months (M24) after rituximab treatment. **e)** Flow cytometry quantification of Treg cell (Foxp3+, CD25high) among peripheral CD4+ in MN patients (n=3) before (D0) and fifteen days (D15) after rituximab treatment. Only 50 patients of the PMMN cohort (out of the 53) were analyzed at baseline due to unavailable PBMC samples. Data are represented as median with interquartile range. P values were determined using two-tailed Mann-Whitney test (**a-c**) or Chi square for trends (**d**).

### Treg levels post-rituximab were associated with chances of remission in MN patients

To explore the effects of rituximab on T helper cell differentiation, we analyzed the proportions of Treg, Th17 and Th1 cells at three, six, nine, 12, 18 and 24 months after rituximab treatment. Enhanced levels of Treg cells were previously reported eight days, three, six or 12 months after rituximab treatment^19–21^. When analyzing the relative proportion of Th1, Th17 and Treg, we observed time-dependent gradual enrichment of Treg cells that reached significance at 24 months post-treatment compared to baseline (p=0.04; Figure 1D). However, we could not distinguish clear differences along the time of follow-up post-rituximab treatment when analyzing separately these three subtypes in the entire cohort (Supplemental figure 5). We then hypothesized that Treg induction might be an early event clearly distinguishable in the first weeks post-rituximab. Indeed, even though analyzed in few patients, we observed a Treg induction when comparing Treg levels among CD4 cells between baseline and two weeks following rituximab administration (Figure 1E).

To determine if an imbalance in T helper cell differentiation is associated with an absence of remission after rituximab treatment, we compared Treg, Th17 and Th1 proportions between patients in remission (REM+) within 24 months after rituximab treatment to those who never entered into remission (REM-). Overall, we observed that, contrary to REM+ patients, the relative proportion of Treg cells tended to decrease over time post-rituximab in REM-patients (Figure 2A-B; Supplemental figure 6). Interestingly, while no difference was detected between both groups of patients at baseline (M0; Figure 2C-D), we showed a significantly higher proportion of Treg cells in REM+ patients compared to REM-patients at six months post-treatment (M6; median 3.20 [2.20; 5.08] vs 1.66 [0.88; 3.48]; p=0.037; Figure 2C). In contrast, no differences of Th17 and Th1 subsets were detected between these groups at M6 (Supplemental figures 6-7). When comparing the relative proportion of Treg, Th17 and Th1, we observed a dysregulated balance of these sub-populations toward lower levels of Treg cells at six months post-Rituximab treatment but not at baseline (M0) in REM-patients compared to REM+ patients (p=0.048; Figure 2E-F). Therefore, low Treg levels six months post-rituximab treatment were associated with poorer chances of remission.

**Figure 2:**
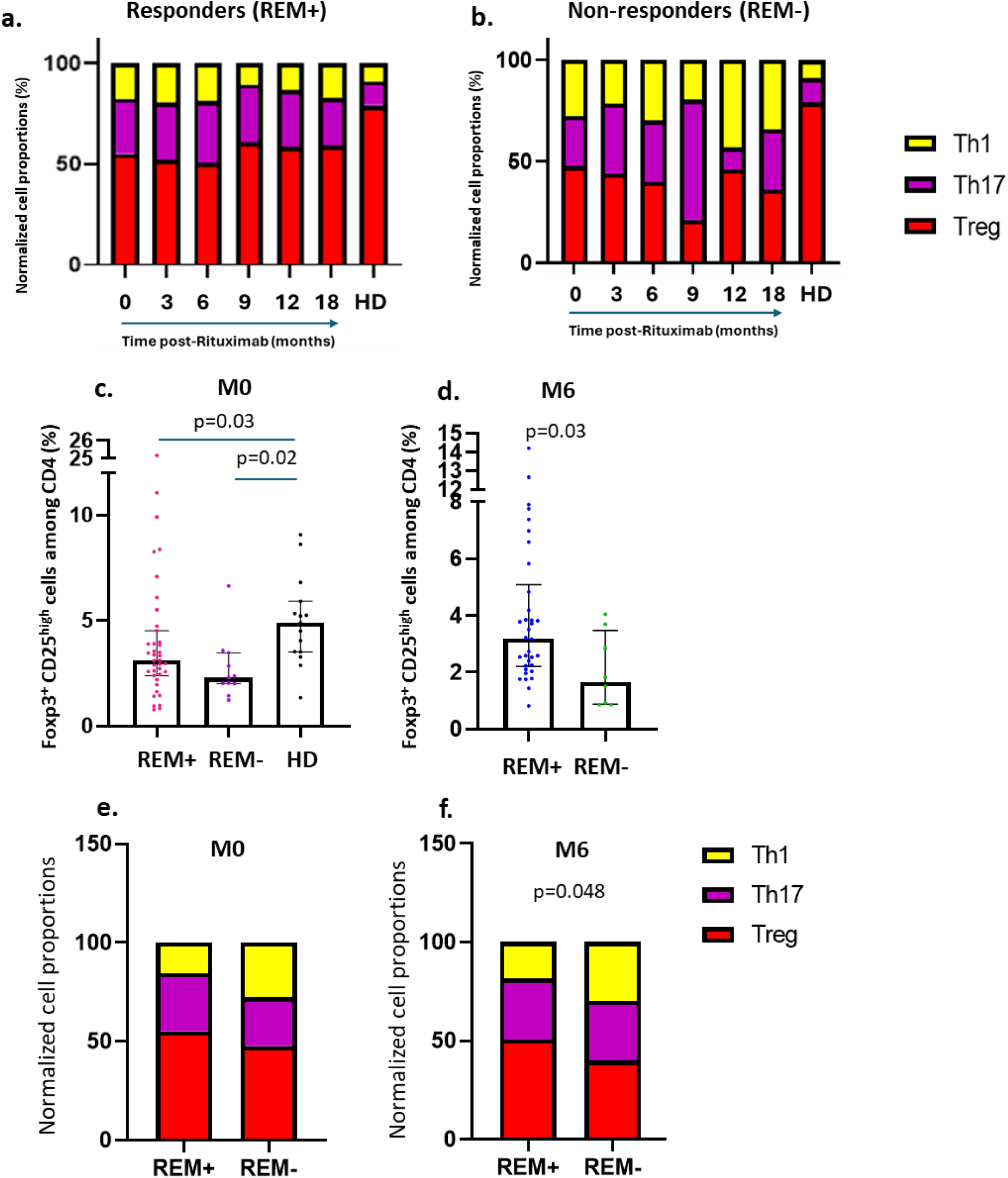
Levels of Treg cells in MN patients are predictive of remission chances after rituximab treatment. MN patients non responding to rituximab treatment have lower levels of Treg cells. **a-b)** Dynamic of the relative proportion of Th1, Th17 and Treg cells after rituximab treatment in either responders **(a)** or non-responders from the PMMN cohort **(b).** The M24 time-point was excluded from this analysis onsidering that only two PBMC REM-samples were available. **c-d)** Flow cytometry quantification of Treg cells (Foxp3+, CD25^high^) among peripheral CD4+ cells before (M0) **(c)** or 6 months (M6) **(d)** after rituximab treatment in MN patients entering (REM+; n=42) or not (REM-; n=11) into remission. e**-f)** Relative proportion of Treg, Th17 and Th1 cells in MN patients entering (REM+; n=42) or not (REM-; n=11) into remission before (M0) **(e)** or 6 months (M6) **(f)** after rituximab treatment. Data are represented as median with interquartile range. P values were determined using either Kruskal Wallis test with Dunn’s post-hoc test **(c)**, two-tailed Mann-Whitney test **(d)** or Chi square for trends **(e-d).**

### Rituximab indirectly induced Treg

Considering the induction of Treg cells by rituximab treatment reported by us (Figure 1E)^19^ and others^20,21^, we sought to determine how this anti-CD20 antibody could act on Th cell differentiation. First, we studied the effects of an *in vitro* rituximab treatment on Th cells of whole blood harvested from MN patients (BIOGEM cohort) and co-stimulated with anti-CD3 and TLR7/8 agonist. Baseline characteristics of these patients were as follows: age: 59.0 ± 14.4; gender: 81% of male; UPCR: 5.4 ± 2.9 g/g; albuminemia: 28.6 ± 4.8 g/L. Twenty hours of rituximab treatment was sufficient to induce Treg cells as observed by cellular increase in Treg levels (CD4+/CD127^low^/CD25^high^ cells; median 42.5 [19.8; 56.4] vs 47.98 [26.17; 61.02]; p=0.048) and plasma IL-10 concentrations after non-specific stimulation (median 874 [609; 1276] vs 939 [685; 1425]; p=0.03; Figure 3 A-B). In line with our previous study^19^, even though lower proportion of CD4+ IL17A+ cells was seen (median 1.75 [0.39; 2.30] vs 0.61 [0.42; 1.03]; p=0.02), no effect of rituximab treatment was detected on plasma IL17A concentrations after non-specific stimulation (median 966 [23.3; 319.5] vs 1188 [19.8; 231.0]; Figure 3C-D). Considering that CD20 expression on T cell was previously reported^23^, we sought to investigate whether rituximab directly influences T cell differentiation. We isolated T cells from peripheral blood samples from healthy donors (N=9) and stimulated them with anti-CD3 and TLR7/8 agonist in presence or absence of rituximab. Strikingly, compared to whole blood cells from the same donors, no Treg induction was detected by flow cytometry after rituximab treatment on isolated T cells (median 0.40 [0.04; 1.56] vs − 0.02 [−0.14; 0.30]; p=0.02; Figure 3E). Similar loss of Treg induction was observed on pooled T and B cells (median 0.10 [0.02; 0.34] vs −0.03 [−0.07; 0.04]; p=0.008; Figure 3F). In contrast, we were not able to detect significant differences on Treg induction by rituximab between whole blood cell and pooled T, B and NK cells (median 0.15 [0.05; 0.43] vs −0.02 [−0.10; 0.16]; p=0.25; Figure 3G). This suggests that rituximab does not directly induce Treg but require at least a cellular environment composed by not only T and B but also NK cells.

**Figure 3:**
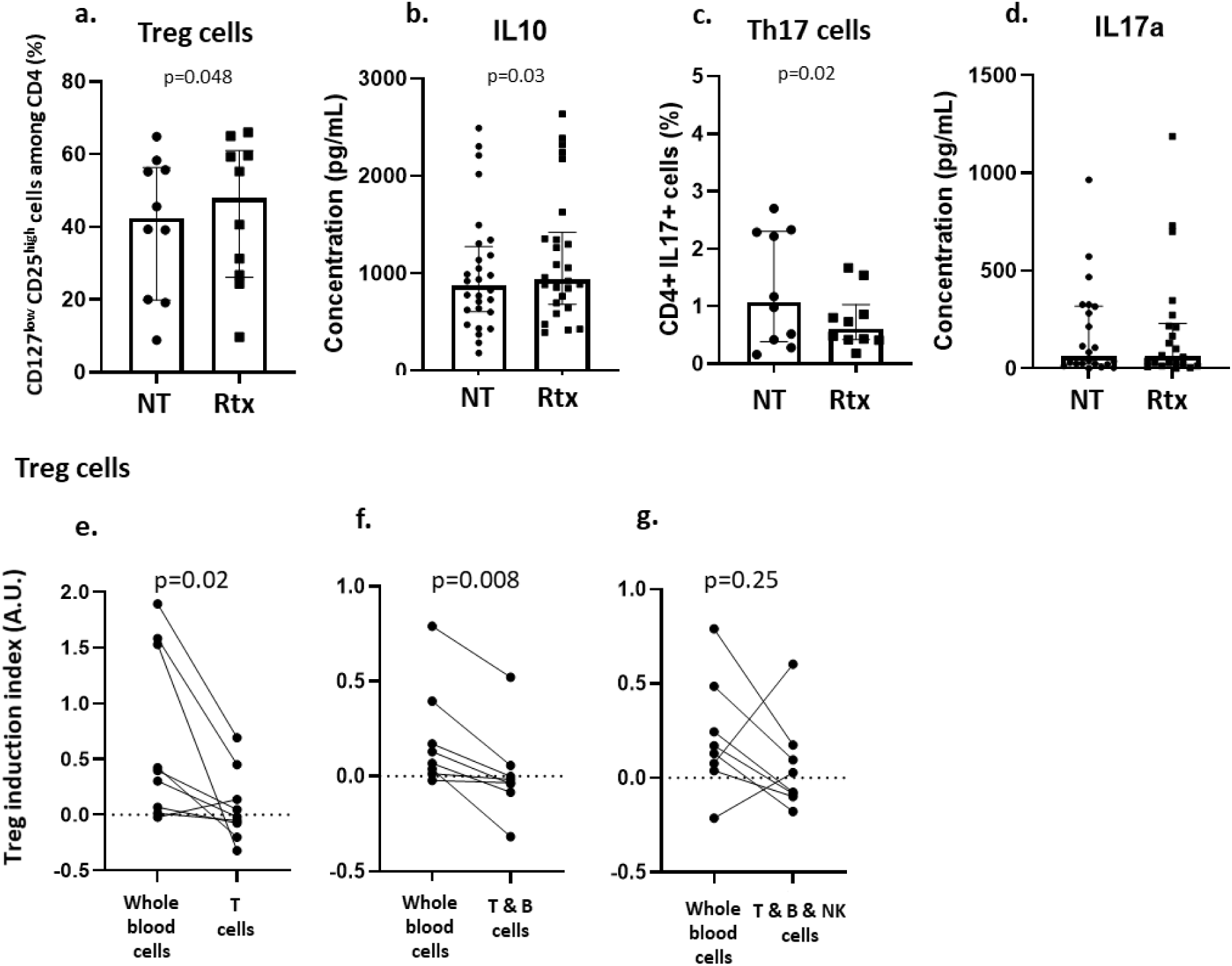
Rituximab indirectly induces Treg cells. *In vitro*, rituximab is able to induce directly Treg on isolated T cells and requires a specific cellular environnement with at least B and NK cells to promote these regulatory cells. **a-d)** Quantification of Treg cells (CD127^low^, CD25^high^) **(a)** or Th17 (IL17+) **(c)** among peripheral CD4+ cells by flow cytometry (n=10) or IL-10 (n=26) **(b)** and IL17a (n=22) **(d)** cytokine levels by ELISA from MN patients’ peripheral blood cell *in vitro* treated (Rtx) or not (NT) by rituximab and after non specific stimulation. **e-g)** Quantification of Treg induction index comparing *in vitro* rituximab treatment of whole blood cells with either isolated T cells **(e)**, B and T cells **(f)** or T, B and NK cells **(g)** from healthy donors (n=8-9) and after non specific stimulation. Treg induction index (TII= (Rtx-NT)/NT) was analysed in all experiments using flow cytometry (CD4+, Foxp3+, CD25high). Data are represented as median with interquartile range. P values were determined using paired Wilcoxon test **(e-g)**. **NT:** non-treated cells; **Rtx:** rituximab treated cells.

### Rituximab-dependent Treg induction required cytokines secreted by activated NK cells

B cell depletion by rituximab binding to CD20 is triggered through direct apoptosis signaling, complement-mediated cytotoxicity (CDC) and antibody-dependent cell-mediated cytotoxicity (ADCC)^24^. NK cells represent the major immune component involved in ADCC. These cells can be subdivided into two major subsets. CD56^brigth^/CD16^dim/-^ NK cells are considered as regulatory cells through their ability to highly secrete cytokines like IFNγ. On the other hand, CD56^dim^/CD16^+^ cells are referred to as the cytotoxic subset of NK cells which releases cytotoxic factors upon activation^25–27^. The CD16a marker (FCγRIIIa), expressed in the latter subset, is an activating receptor involved in ADCC by binding to the Fc portion of IgG antibodies^27^. This interaction is crucial for ADCC-dependent B cell depletion by anti-CD20 antibodies^24^. In order to provide further mechanistic insights explaining how NK cells could mediate rituximab-dependent Treg induction, we performed complementation assays to assess for a mechanism involving cytokine signaling. Complemented media from isolated B cells or pooled B and NK cells treated with rituximab were transferred onto isolated T cells prior to being stimulated with anti-CD3 and TLR7/8 agonist. Strikingly, while no Treg induction was observed for T cells incubated with B cell-derived complemented medium, we showed that complemented medium derived from pooled B and NK cells pre-treated with rituximab was sufficient to induce Treg (mean 0.04 ± 0.02 vs 0.33 ± 0.12; p=0.048; Figure 4A). This suggests that Treg induction by rituximab is mediated by soluble factors released in the medium by activated NK cells during their interaction with B cells and rituximab.

**Figure 4:**
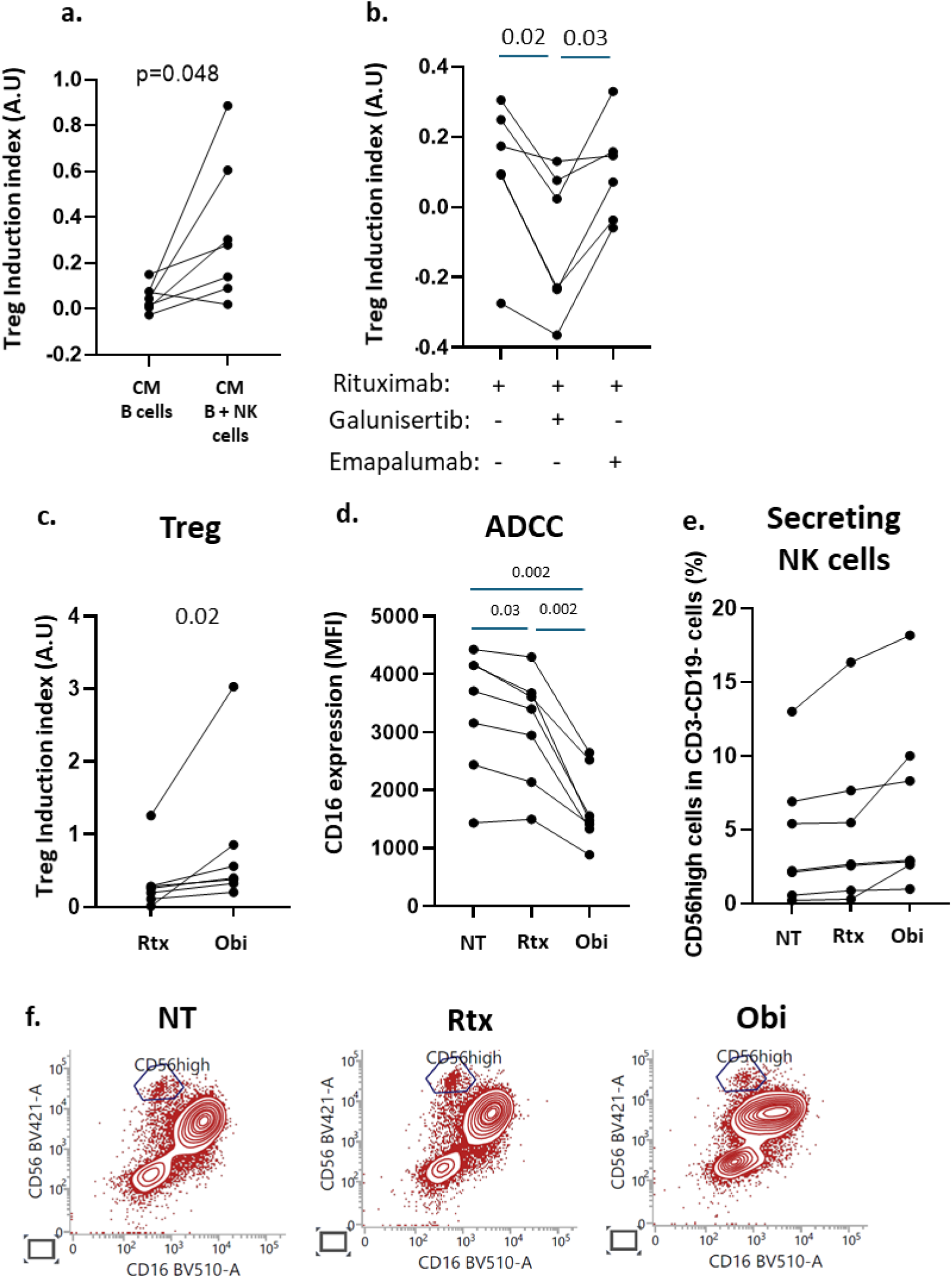
Rituximab induces Treg cells through cytokine release by activated NK cells during ADCC. *In vitro*, Treg induction by rituximab relied on cytokine factors secreted by NK cells during ADCC-mediated B cell depletion. **a)** Quantification of Treg induction index comparing treatment of isolated T cells from healthy donors (N=7) with complemented medium derived from B cells (CM B cells) or a pool of B and NK cells (CM B + NK cells) treated or not with rituximab. **b)** Quantification of Treg induction index comparing treatment of isoled T cells with complemented medium pre-treated with galunisertib or emapalumab and derived from a pool of B and NK cells treated with rituximab (N=6). **c-e)** Quantification of Treg induction index **(c)** CD16 expression levels of NK cells (CD45+CD3-CD19-) **(d)** or CD56^high^CD16^dim/-^ **(e)** using flow cytometry comparing treatment of peripheral blood samples of healthy donors (N=7) with rituximab or obinutuzumab after non specific stimulation. **f)** Representative CD56/CD16 flow cytometry plot of peripheral blood samples treated with rituximab or obinutuzumab after non specific stimulation. Treg induction index (TII= (Rtx-NT)/NT) was analysed in all experiments using flow cytometry (CD4+, Foxp3+, CD25high). P values were determined using paired T-test **(a)**, paired Wilcoxon test **(c)** or paired RM one-way ANOVA test with Tukey’s post-hoc correction **(b; d-e)**. **NT:** non-treated cells; **Rtx:** Rituximab treated cells; **Obi:** Obinutuzumab; **MFI:** mean fluorescence intensity

Beside IFNγ, even though barely reported and characterized, NK cells have been described to secrete TGFβ, a potent Treg inducer^28,29^. To investigate how NK cells mediate rituximab-dependent Treg induction, we then focused on TGFβ and IFNγ and tested whether inhibiting these factors would impede Treg induction. We showed that adding TGFβ receptor inhibitor (galunisertib) to complemented medium derived from rituximab-treated B and NK cells abolished the Treg induction seen in figure 4A (mean: Rtx alone: 0.13 ± 0.08 VS Rtx + galunisertib: −0.1 ± 0.08; p=0.02; Figure 4B). However, the anti-IFNγ antibody (emapalumab) had no inhibitory effects. Therefore, Treg induction by rituximab seems to be mediated by cytokines (mainly TGFβ) produced by NK cells, probably during the mechanism of B cell depletion by ADCC. In line with this, treating peripheral blood samples with the next generation of anti-CD20 antibody, obinutuzumab, which was glycoengineered to enhance its ADCC activity^30^, resulted in higher Treg induction compared to rituximab (median: Rtx: 0.26 [0.11; 0.29] vs Obi: 0.4 [0.33; 0.86]; p=0.02; Figure 4C). Moreover, CD16a down-regulation has been described following NK cell activation during ADCC to favor immune cell detachment^31,32^. By analyzing CD16 mean fluorescence intensity, we observed that rituximab treatment of peripheral blood samples leads to a significant CD16a shedding (mean: 3307 ± 1091 vs 3081 ± 968.2; p= 0.03; Figure 4D). Notably, we noticed higher CD16a down-regulation when treating blood samples with obinutuzumab compared to rituximab reflecting enhanced ADCC activity of this second generation anti-CD20 antibody (1691 ± 648.3 vs 3081 ± 968.2; p=0.002; Figure 4D and 4F). However, no difference was detected when comparing the proportions of CD56^brigth^/CD16^dim/-^ cells between untreated samples, rituximab- or obitunuzumab-treated peripheral blood samples (Figure 4E-F). Treg induction cannot be therefore explained by increased CD56^brigth^/CD16^dim/-^ regulatory cell number. Overall, these results show that Treg induction by rituximab is mediated by cytokines secreted by activated NK cells probably during ADCC-mediated B cell depletion.

## DISCUSSION

Even though MN is triggered by the production of auto-antibodies targeting podocytes, Th cell phenotype imbalance and consequent loss of immune tolerance is suggested to play an important role in MN onset and progression^19–21,33^. While Th17 lymphocytes are pro-inflammatory and promote auto-immunity, Treg cells counteract auto-immunity by ensuring immune tolerance and homeostasis. Disturbed Th17/Treg balance skewed toward the pro-inflammatory pathway is highlighted in auto-immune disorders^13,14^ and mouse studies demonstrated that inhibiting Th17 or inducing Treg cells can attenuate auto-immune disease onset and progression^34–36^. Here, using a randomized multicentric prospective trial, we confirmed that MN patients at baseline are affected by a disbalance of the Th17/Treg ratio with increased prevalence of Th17 cells and defects in Treg cell proportions. Interventions aiming at counteracting this imbalance would therefore rise as interesting strategies to improve MN patient health.

Anti-CD20 therapy in auto-immune diseases, such as rituximab treatment, has been developed to deplete B lymphocytes and therefore reduce circulating pathogenic auto-antibodies^10,24^. Increasing interest has emerged into deciphering the impact of anti-CD20 antibodies on T cell response in MN patients. Rituximab has indeed the ability to dampen T cell activation and favors Treg induction in multiple auto-immune diseases such as systemic lupus erythematosus, mixed cryoglobulinaemia vasculitis, refractory myasthenia gravis or mixed cryoglobulinemia vasculitis^37–43^. Although rituximab is the first line treatment in MN^9^, limited studies attempted to characterize its impact on Th cells. Most of them described an enrichment in Treg cells after rituximab treatment^19–21,44^ whereas Fervenza et al. were not able to detect any effect on this subtype^12^. In the present work, while no differences were detected for Th17 and Th1 subsets, we showed a gradual increase in Treg cell relative proportion following rituximab treatment. Consistent with previous studies^20,21^, we also highlighted that MN patients in remission have higher levels of Treg cells following rituximab treatment compared to patients that do not achieve remission. This would suggest that Treg induction by rituximab could favor remission.

Despite evidence of enhanced Treg cell levels following rituximab treatment in auto-immune diseases, the mechanism behind this induction remains unclear. Considering that rituximab is able to deplete CD3^+^CD20^dim^ cells^45,46^, a direct effect of this anti-CD20 on T cells was previously suggested to explain this phenomenon. By treating isolated T cells with rituximab, we demonstrated that rituximab does not directly induce Treg cells. Moreover, Treg induction was also lost when treating isolated T and B cells, excluding the involvement of B cell repressive functions^47,48^. Notably, we revealed that Treg induction by rituximab requires a specific cellular environment including at least B and NK cells as observed by treating pooled T, B and NK cells. As shown in our complementation assays with isolated B and NK cells, Treg induction relies on cytokine production, and inhibiting TGFβ signaling abolished this phenomenon. This suggests that rituximab-dependent Treg induction is mediated by cytokines, mainly TGFβ, probably produced by activated NK cells during B cell depletion by ADCC. In line with this, we showed that increasing ADCC-dependent B depletion by using the second generation of anti-CD20 antibody, obinutuzumab^30,49,50^, leads to higher enrichment of Treg cells. Increase in TGFβ mRNA or serum levels after rituximab treatment has previously been reported in, respectively, SLE and Non-Hodgkin’s Lymphoma patients^38,51^. Here, we did not assess whether patients in remission are associated with higher serum levels of TGFβ. Considering that high Treg levels are associated with better chances of remission, additional studies on TGFβ serum levels following rituximab treatment in MN patients could determine whether this cytokine could be a good predictor of clinical outcome.

In this study, we unraveled that NK cell activation is central for rituximab-dependent Treg induction. Defects in CD56+ NK cell population were previously described in MN^20^. Enrichment in NK cells also occurred following rituximab treatment and higher levels of NK cells are associated with better chances of remission in MN patients^20,52^. The Fc receptor CD16a (FCγRIIIa) is essential for IgG dependent NK cell activation. One main functional polymorphism of the FCGR3A (encoding CD16a) has been extensively studied, where CD16a-bearing 158V phenotype has better affinity to IgG and promote higher ADCC compared to the 158F phenotype^27^. The low affinity variant is associated with susceptibility to develop rheumatoid arthritis and systemic lupus erythematosus whereas the high affinity phenotype confers a better clinical outcome following rituximab treatment^53–56^. Such polymorphism has never been studied in MN patients. The results of our current study would then pave the way to investigations relating NK cell activation, CD16a variants and Treg with clinical outcome upon anti-CD20 treatment of MN patients.

Our present work suggests that enhancing anti-CD20 antibody potency to induce ADCC favors regulatory T cells. While rituximab mainly relies on CDC, obinutuzumab was glycoengineered to enhance ADCC activity of anti-CD20 antibodies^30,49^. Recent studies point out that obinutuzumab is more efficient in treating rituximab-resistant MN patients or as a first-line treatment^57–59^. However, *in vivo* effects of obinutuzumab on Treg levels remain to be determined in MN and other auto-immune diseases. Investigating whether higher efficiency of obinutuzumab solely stem from better B-cell depletion or if it relies on reorienting T cell response toward regulatory cells would open to new strategies to improve clinical outcome of MN patient treatment.

The main limitations of the study include the relative limited number of patients enrolled in the PMMN cohort which only allowed to analyze 11 non-responders. Moreover, due to multi-centric shipment constrains, some PBMC could not be isolated leading a relative variability in the number of samples analyzed for each time point. However, this study relies on a powerful multi-center and randomized design of the PMMN cohort with a 24-month follow-up. This was combined with *in vitro* experiments on fresh human samples that provided mechanistic insights on rituximab-mediated Treg induction.

In conclusion, *in vivo* induction of regulatory T cell by rituximab is associated with better chances of remission. Moreover, Treg induction by anti-CD20 antibodies such as rituximab or obinutuzumab relies on cytokines released by activated NK cells, probably during ADCC-dependent B-cell depletion. Development of novel therapeutic strategies enhancing NK cell activation and ADCC could then be beneficial for MN patients by favoring immune tolerance.

## Contributors

BS-P and MEM contributed to the conceptualization of the study. VB, KZ and EV-S performed data curation. VB, YC and MEM were responsible for the formal analysis. BS-P, CF, MC and MT were in charge of funding acquisition. VB; YC; EV-S; SNC; SA and MEM performed the investigations.BS-P, MC and MT were in charge of clinical data and biosamples collection. BS-P provided senior mentorship and supervision. BS-P and MEM wrote the original draft. All authors read and approved the final version of the manuscript.

## Data Availability

The data that support the findings of this study are available from the corresponding author upon reasonable request.

## Acknowledgments

We thank all the medical, paramedical and technical staff implicated in patient care and laboratory analyses, both from the study coordinator site and from the centers who participated in the study. This work was sponsored by the Centre Hospitalier Universitaire de Nice (University Hospital of Nice) for regulatory and ethic submission. This work was supported as part of the national plan for rare diseases by the French Ministry of Health and also by a grant PHRC: Clinical Research Hospital Program from the French Ministry of Health (PHRC National 2017).

## Supplemental Material

This article contains the following supplemental material:

**Supplemental figure 1:** Flow chart for PMMN cohort analysis.

**Supplemental figure 2:** Flow cytometry gating strategy for PMMN cohort analysis.

**Supplemental figure 3:** Flow cytometry gating strategy for BIOGEM cohort analysis.

**Supplemental figure 4:** Flow cytometry gating strategy for Treg and NK cell analysis on healthy donors.

**Supplemental figure 5:** Analysis of Th cell subset in MN patients at baseline and after rituximab treatment.

**Supplemental figure 6:** T cell subset kinetics after rituximab treatment.

**Supplemental figure 7:** Levels of Th17 and Th1 cells at baseline and 6 months post-rituximab treatment relative to the clinical outcome.

