## Supplemental Material for "Rituximab counteracts loss of tolerance in membranous nephropathy patients through NK-mediated Treg induction"

**Mounir El Maï[1,2,3], Maxime Teisseyre[1,2,3], Yousra Cheddadi[2], Vesna Brglez[1,2,3], Erendira Vazquez-Salazar[1,2,3], Sarah Nahon Carzo[2], Sami Addou[2], Kevin Zorzi[1,2,3], Marion Cremoni[2,3], Céline Fernandez[1,2,3], Barbara Seitz-Polski[1,2,3]**

### Affiliations:

[1]Centre Hospitalier Universitaire de Nice, Centre de Référence Maladies Rares Syndrome Néphrotique Idiopathique et Glomérulonéphirte Extra-Membraneuse, Nice, France
[2]Centre Hospitalier Universitaire de Nice, Laboratoire d'Immunologie et de Thérapie Cellulaire, Nice, France
[3]Université Côte d'Azur, CNRS, INSERM, IRCAN, Nice, France

### Table of contents for Supplemental Material

**Supplemental figure 1:** Flow chart for PMMN cohort analysis …………………………………………………..2

**Supplemental figure 2:** Flow cytometry gating strategy for PMMN cohort analysis ……………………………..3

**Supplemental figure 3:** Flow cytometry gating strategy for BIOGEM cohort analysis …………………………..4

**Supplemental figure 4:** Flow cytometry gating strategy for Treg and NK cell analysis on healthy donors .......5

**Supplemental figure 5:** Analysis of Th cell subset in MN patients at baseline and after rituximab treatment ..6

**Supplemental figure 6:** T cell subset kinetics after rituximab treatment …………………………………………7

**Supplemental figure 7:** Levels of Th17 and Th1 cells at baseline and 6 months post-rituximab treatment relative to the clinical outcome …………………………………………………………………………………….8

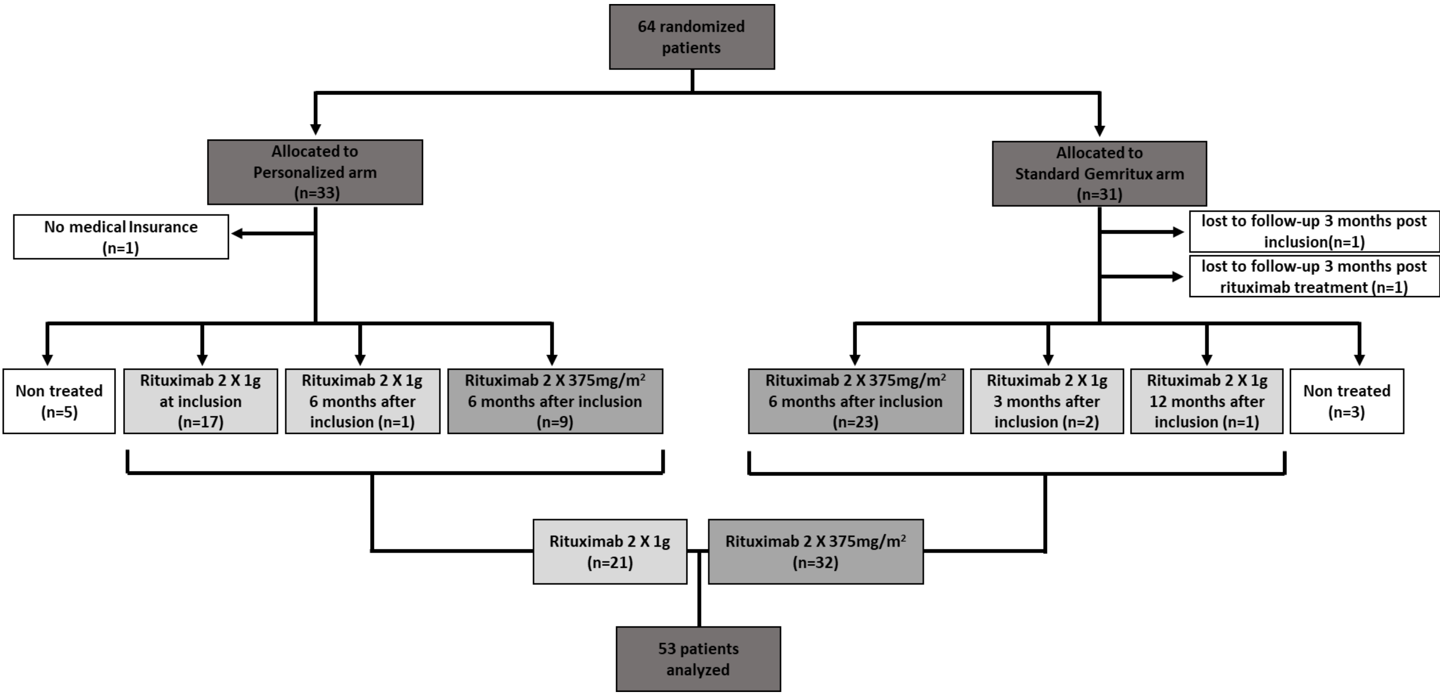


**Supplemental figure 1: Flow chart for PMMN cohort analysis.**

Among the 64 randomized patients initially included in the PMMN study, T helper cell populations were analyzed for 53 patients from both the personalized arm (N=27) and the standard Gemritux arm (N=26). The remaining 11 patients were excluded from this ancillary study since they either did not receive rituximab during the course of the study (N=8), due to loss to follow-up (N=2) or because of the absence of medical insurance (N=1).

In the Personalized arm, 17 patients were treated with 2x1g of rituximab at two weeks interval directly after inclusion while nine and one patients received, after 6 months of non-immunosuppressive antiproteinuric treatment (NIAT), respectively two 375 mg/m2 rituximab injections (at one-week interval) or two 1g rituximab injections (at two weeks interval). The remaining six patients that had either no rituximab treatment (N=5) or no medical insurance were not analyzed in this ancillary study.

In the standard Gemritux arm, 23 patients received NIAT for 6 months before being treated with two 375 mg/m2 rituximab injections (at one-week interval). Three patients deviated from the standard treatment and received two high doses of rituximab (2x1g at two weeks interval) with after either 3 months (N=2) or 12 months (N=1) of NIAT. The remaining five patients that had either no rituximab treatment (N=3) or lost to follow-up at inclusion or 3 months after rituximab treatment were not analyzed in this ancillary study.

Overall, 32 patients who received two 375 mg/m2 rituximab injections (at one-week interval) and 21 patients who received two 1g rituximab injections (at two weeks interval) were analyzed in the ancillary study.

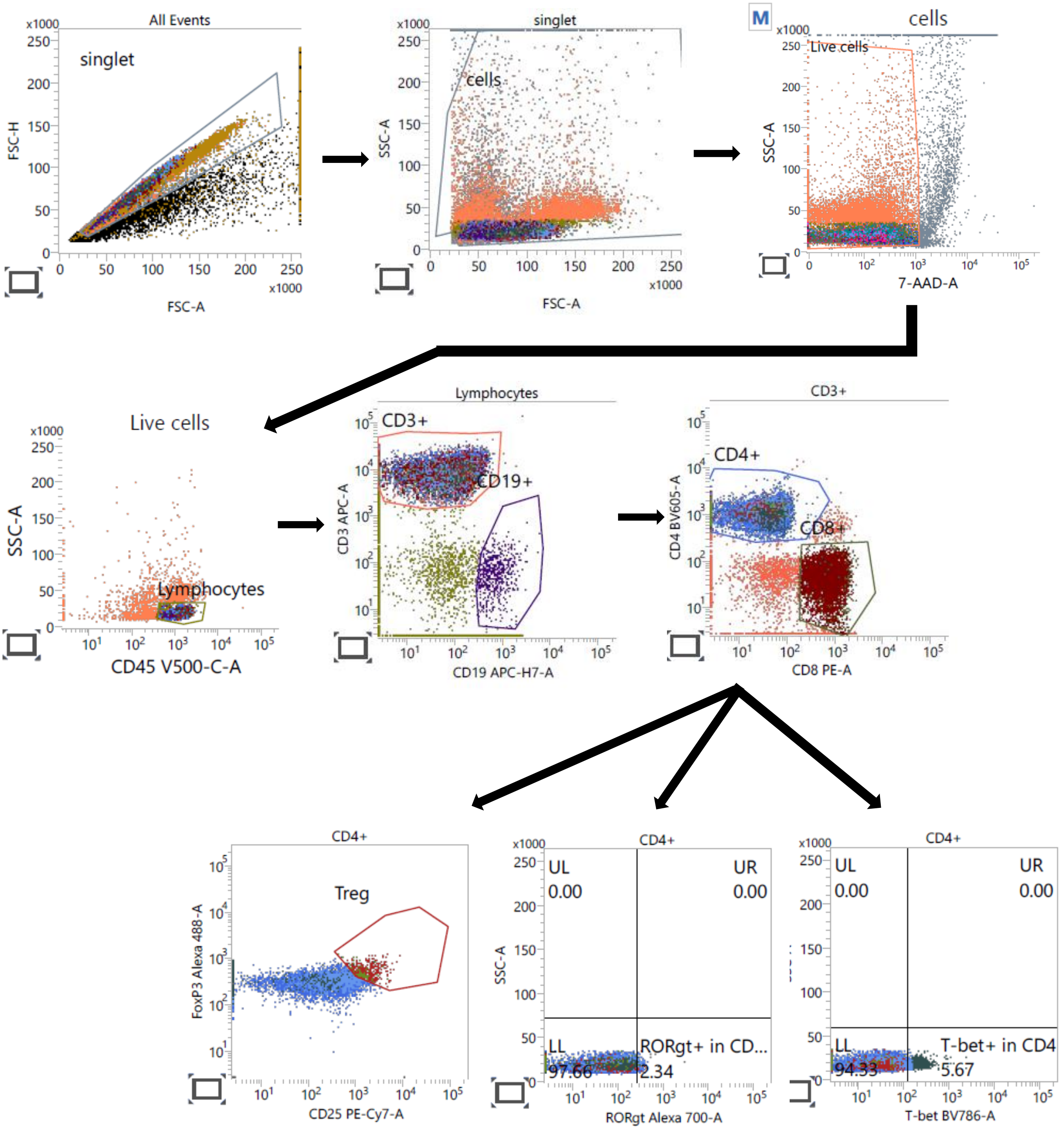


**Supplemental figure 2: Flow cytometry gating strategy for PMMN cohort analysis.** Peripheral blood mononuclear cells of the PMMN cohort were analyzed by flow cytometry. Single cell population cells were separated for size and granularity and dead cells excluded using 7-AAD. Lymphocytes were localized by CD45 expression. Treg cell (Foxp3+, CD25high), Th17 cells (RORgt+) and Th1 cells (T-bet+) were determined within CD45+CD3+CD4+ as depicted in this figure.

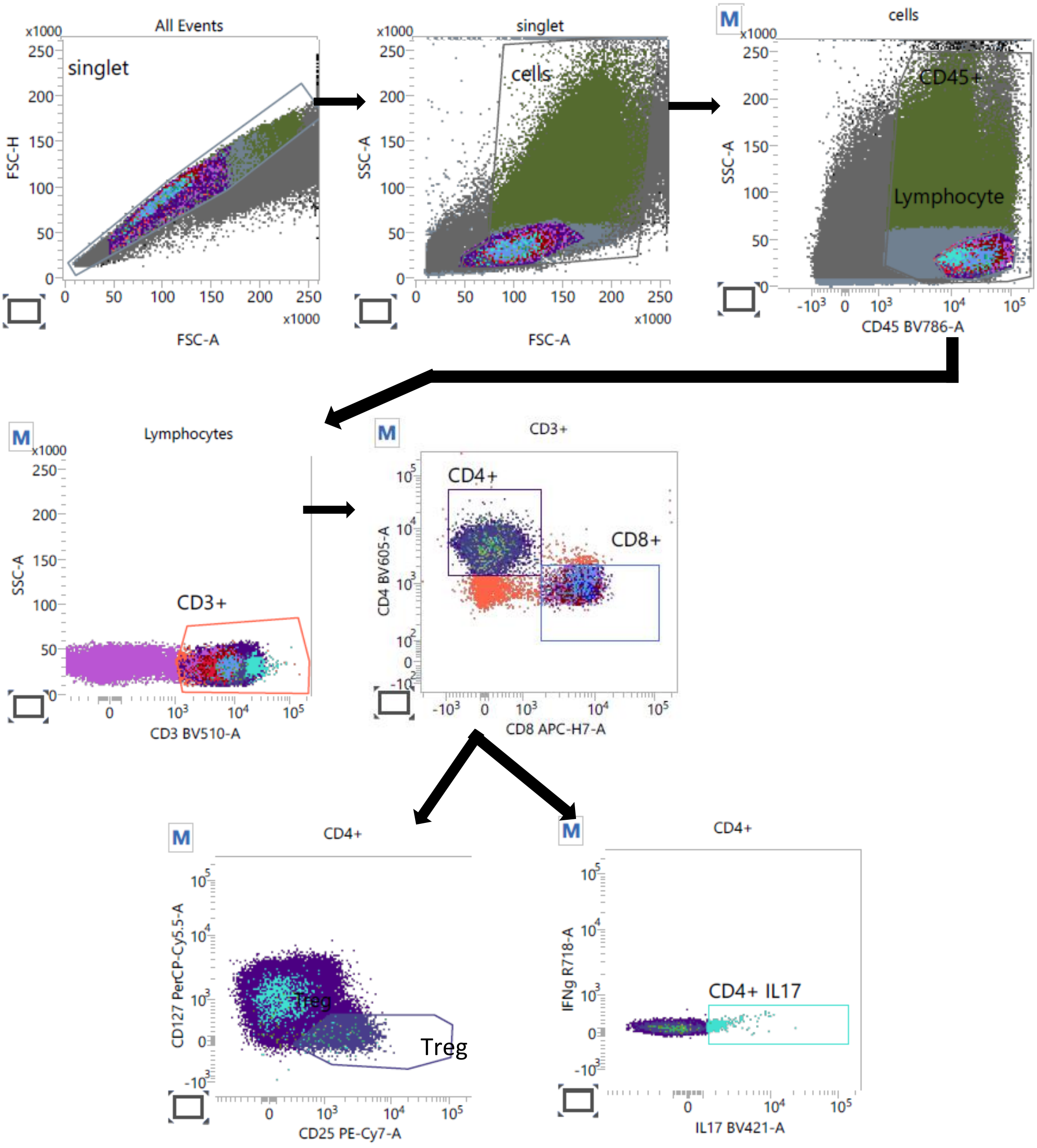


**Supplemental figure 3: Flow cytometry gating strategy for BIOGEM cohort analysis.** Single cell population was determined by FSC-A/FSC-H and used for analysis and then separated for size and granularity. Lymphocytes were localized by CD45 expression. Treg cell (Foxp3+, CD25high) were determined within CD45+CD3+CD4+ as depicted in this figure. Subpopulation expressing CD16 and CD56 were determined within CD3-CD19- cells.

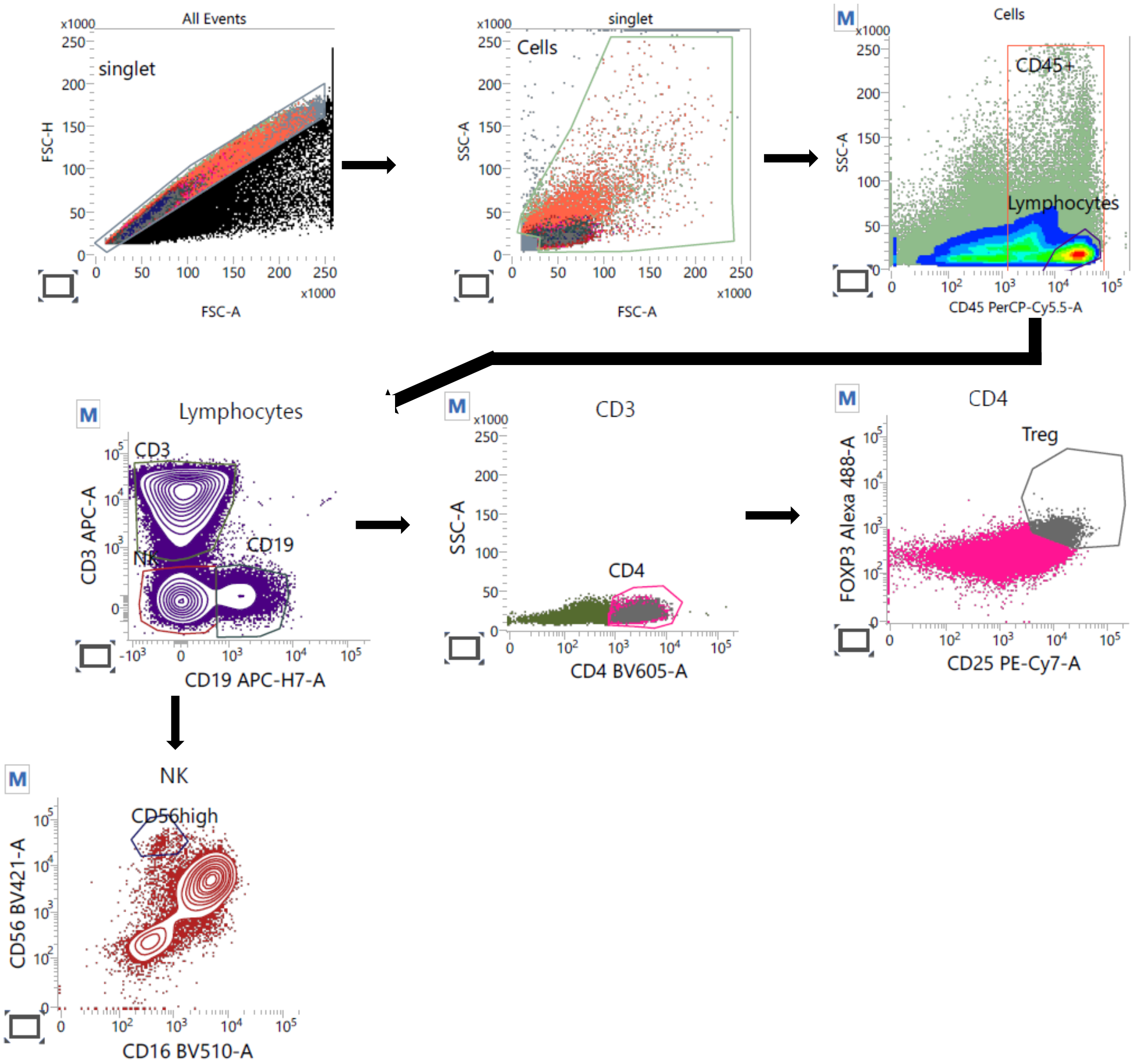


**Supplemental figure 4: Flow cytometry gating strategy for Treg and NK cell analysis on healthy donors.** Single cell population were determined by FSC-A/FSC-H and used for analysis and then separated for size and granularity. Lymphocytes were localized by CD45 expression. Treg cell (Foxp3+, CD25high) were determined within CD45+CD3+CD4+ as depicted in this figure. Subpopulation expressing CD16 and CD56 were determined within CD3-CD19- cells.

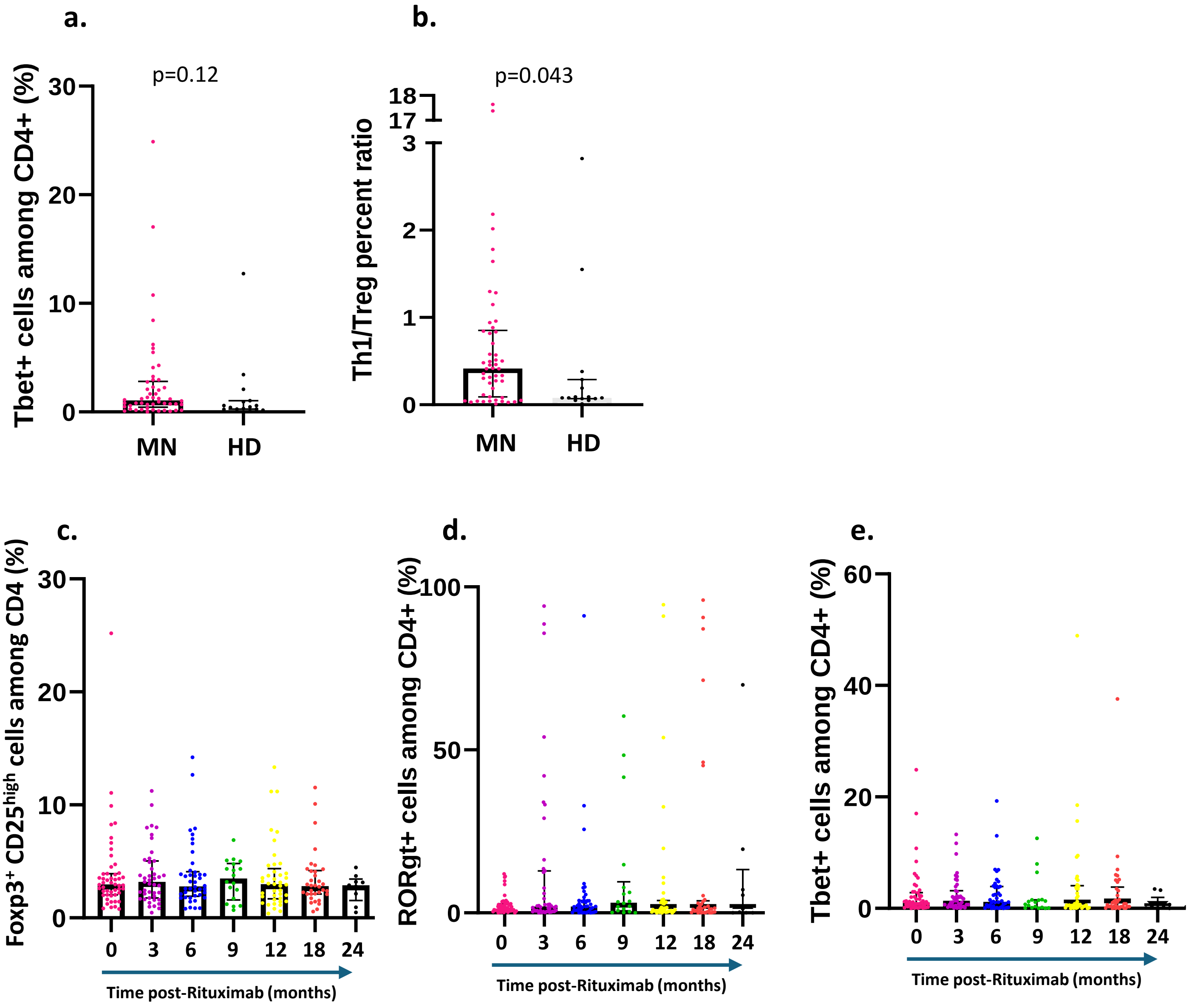


**Supplemental figure 5: Analysis of Th cell subset in MN patients at baseline and after rituximab treatment. a-b)** Flow cytometry quantification of Th1 (T-bet+) among peripheral CD4+ cells (**a**), Th1/Treg ratio **(b)** at baseline in MN patients from the PMMN cohort (MN; n=50) compared to healthy donors (HD; n=15). **c-d)** Flow cytometry quantification of Treg cell (Foxp3+, CD25high) **(c),** Th17 cells (RORgt+) **(d)** and Th1 cells (T-bet+) **(e)** among peripheral CD4+ cells after rituximab treatment in MN patients. Only 50 patients of the PMMN cohort (out of the 53) were analyzed at baseline due to unavailable PBMC samples. Data are represented as median with interquartile range. P values were determined using two-tailed Mann-Whitney test.

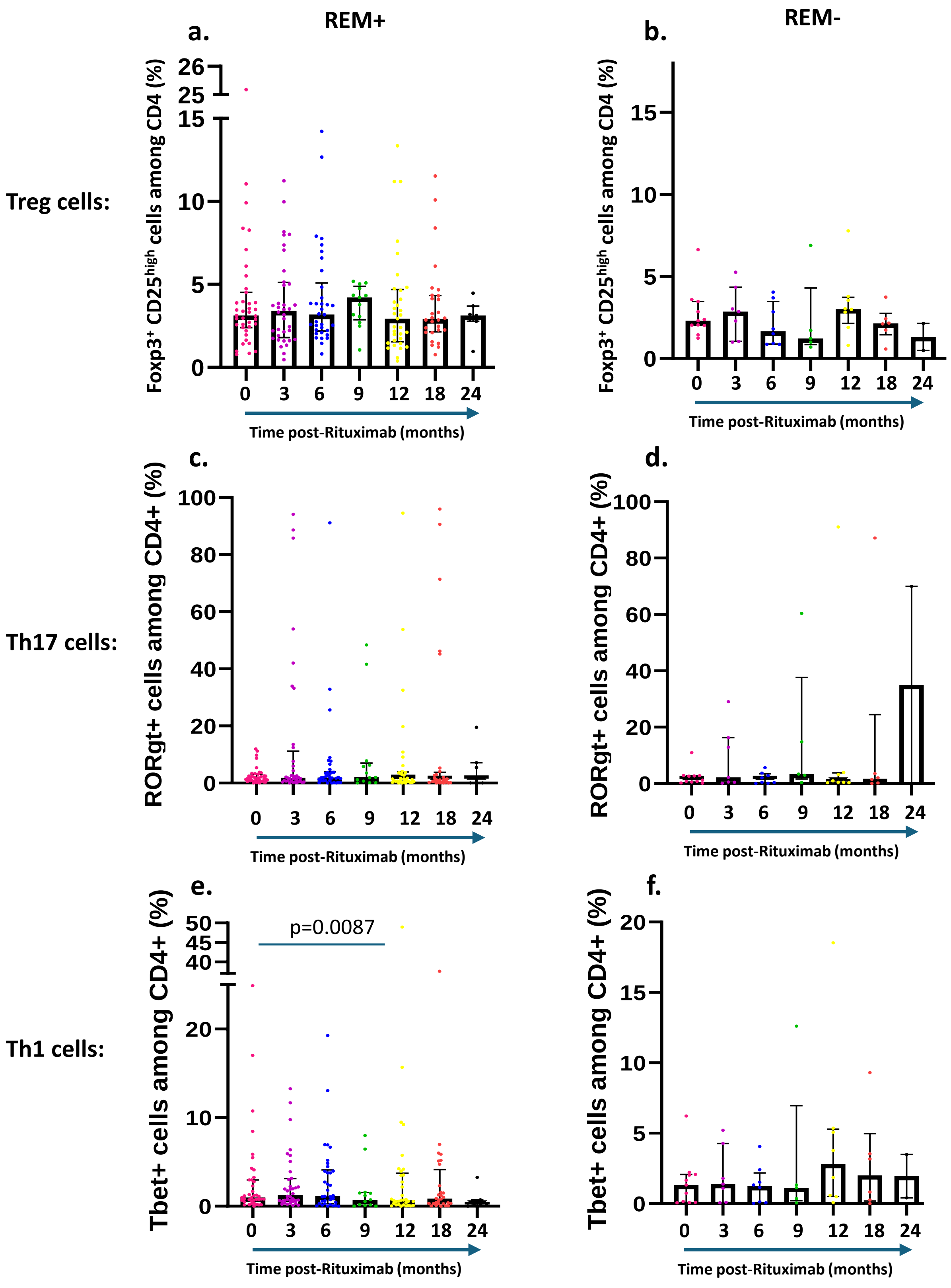


**Supplemental figure 6: T cell subset kinetics after rituximab treatment. a-f)** Flow cytometry quantification of Treg cell (Foxp3+, CD25high) **(a-b)**, Th17 cells (RORgt+) **(C-D)** and Th1 cells (T-bet+) **(e-f)** among peripheral CD4+ cells after rituximab treatment in MN patients from the PMMN cohort entering (REM+; n=42; **a-c-e**) or not (REM-; n=11; **b-d-f**) into remission. Data are represented as median with interquartile range. P values were determined using paired Wilcoxon test.

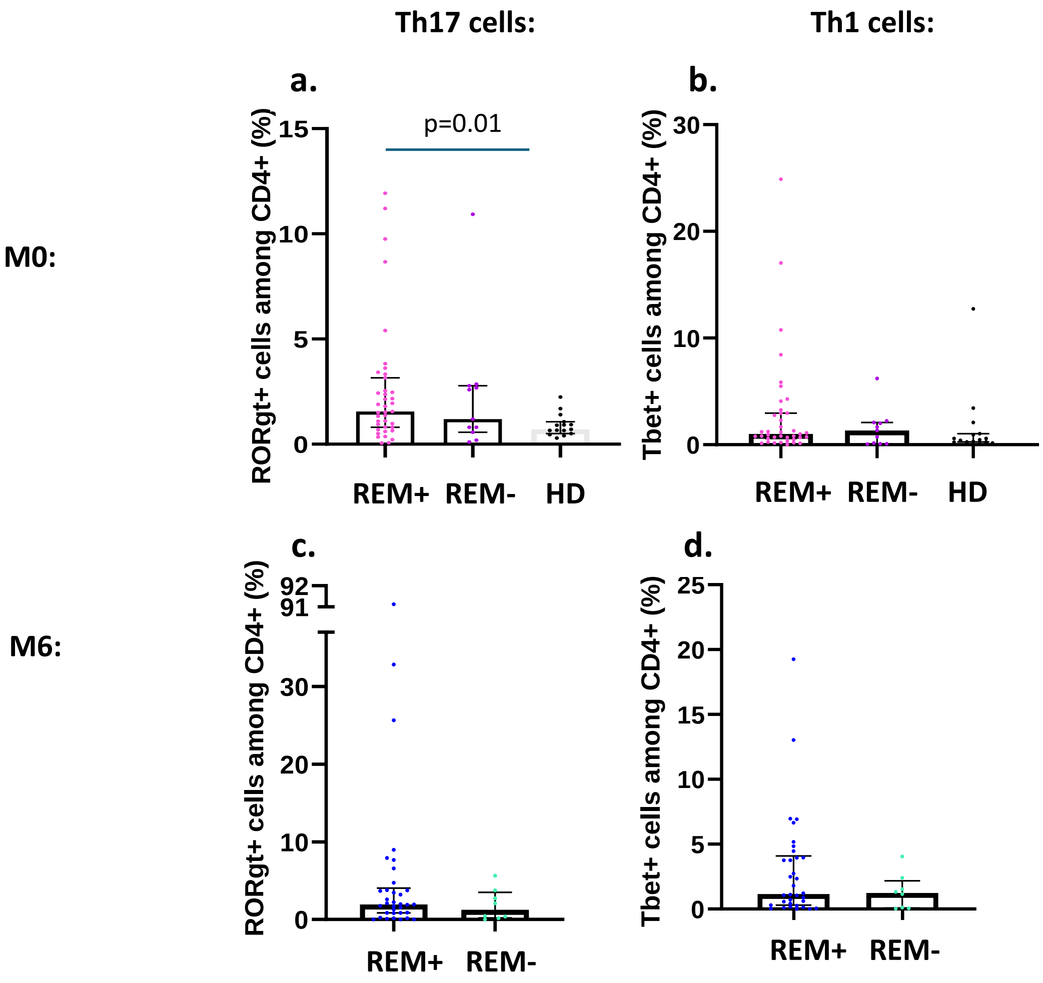


**Supplemental figure 7: Levels of Th17 and Th1 cells at baseline and 6 months post-rituximab treatment relative to the clinical outcome. a-d)** Flow cytometry quantification of Th17 cells (RORgt+) **(a; c)** and Th1 cells (T-bet+) **(b; d)** among peripheral CD4+ cells before (M0) **(a-b)** or 6 months (M6) **(c-d)** after rituximab treatment in MN patients from the PMMN cohort entering (REM+; n=42) or not (REM-; n=11) into remission. Only 50 patients of the PMMN cohort (out of the 53) were analyzed at baseline due to unavailable PBMC samples. Data are represented as median with interquartile range. P values were determined using either Kruskal Wallis test with Dunn's post-hoc test **(a-b) or** two-tailed Mann-Whitney test **(c-d)**.
